# Impact of the COVID-19 pandemic on tuberculosis notification in Brazil

**DOI:** 10.1101/2022.09.05.22279616

**Authors:** Daniele M Pelissari, Patricia Bartholomay, Fernanda Dockhorn Costa Johansen, Fredi A Diaz-Quijano

## Abstract

**Background:** The COVID-19 pandemic notably impacted tuberculosis notification and detection in Brazil. We estimated the number of unnotified tuberculosis cases by group population over the first two years (2020-2021) of the pandemic.

**Methods:** We extracted tuberculosis case notifications from routine national surveillance records and population from Ministry of Health. We estimated trends for case notification during pre-pandemic period (2015–2019), stratified by sex, age group, and State with a mixed-effects model. We calculated the unnotified cases during 2020-2021 as the difference between expected, and reported values.

**Results:** We estimated 11647 (95% uncertain interval [95%UI]: 829,22466) unnotified cases for 2020; and, 6170 (95%UI: -4629,16968) for 2021; amounting 17817 unnotified cases over the two years. Of the estimated expected tuberculosis cases in 2020 and 2021, 11.2% were not notified. Across sex and age, men aged 30-59 years had the highest number of unnotified cases, and men aged 0-14 years had the highest proportion of unnotified cases. Case underreporting was significant for 13 (of the 27 States) in 2020, and for four in 2021.

**Conclusions:** Tuberculosis cases notification decreased substantially during the COVID-19 pandemic in Brazil. Our analysis helped identify the most affected populations to plan strategies to mitigate the effects of the pandemic on tuberculosis control.

**Research in context:** *Evidence before this study:* A systematic review was conducted to retrieve studies that aimed the impact of the COVID-19 pandemic on tuberculosis detection in PubMed with the following terms: “(TB or tuberculosis) and (incidence or case or notification or burden) and (COVID-19 or pandemic)” from January 2020 to May 2022, returning 189 records. Out of these studies, we analyzed 17 that reported a decrease in tuberculosis notification during the pandemic years, and most of them with data only from the first year of the pandemic. Two studies were carried out with Brazilian data. One of them focused on the number of tuberculosis consultations at the benning of the pandemic, and the other was a government bulletin describing tuberculosis notification. As far as we know, no study has examined the tuberculosis case notification in Brazil during the two years of the pandemic, by group population. Furthermore, none of them had predicted the expected cases considering local trends in both the incidence of tuberculosis and its main determinants.

*Added value of this study:* Using tuberculosis case reports from routine national surveillance registries, we estimated case notification trends during the pre-pandemic period (2015–2019), stratified by sex, age group, and State and calculated the unnotified cases during 2020-2021. Brazil lost 11647 (95% uncertain interval [95%UI]: 829,22466) tuberculosis cases in 2020; and, 6170 (95%UI: - 4629,16968) in 2021, which represents 11.2% of underreporting in both years. Across sex and age, men aged 30 to 59 years had the highest number of unnotified cases, and men aged 0 to 14 years had the highest proportion of unnotified cases. Case underreporting was significant for 13 (of the 27 States) in 2020, and for four in 2021.

*Implications of all the available evidence:* The COVID-19 pandemic had a catastrophic effect in tuberculosis notification in Brazil during 2020 and 2021. This resulted in a setback in progress made over decades in tuberculosis control, and highlight the threat posed by tuberculosis transmission. Several lessons learned from response to COVID-19 provide an opportunity to improve the notification of respiratory diseases.

## Introduction

In the last two years, the world’s attention has been focused on the most lethal pandemic seen for over a century.^1^ Resources of surveillance programs that had been structured and organized for decades moved to the COVID-19 response. This had an overwhelming effect on global health, and tuberculosis (TB) services have been extremely affected.^2^ Community-based screening, contact tracing, and active case finding were discontinued.^1,2^ Number of patients initiating treatment declined precipitously, and preventive therapy enrolment and care-seeking initiatives were compromised.^1–3^

Before the pandemic, Brazil was known to have a decentralized TB care network, which placed it in the world ranking of countries with the highest TB case detection rates.^4^ However, like many other countries, it was affected by COVID-19 in early 2020 (first case reported on 26 February 2020), and had to deal with three waves, being the second one the deadliest, and the Omicron-dominated wave the most transmissible one.^5^ Since the beginning of the pandemic, there have been concerns over the effects to the routine TB healthcare service, which might lead to cutbacks in the progress made so far on TB case detection.^6^

The World Health Organization (WHO) assessed that the COVID-19 pandemic has led to a substantial decrease in the number of individuals newly diagnosed with TB and reported to authorities. It estimated that 10 million people developed TB in 2020, but only 5.8 million cases were diagnosed and reported, which represents a global drop between 2019 and 2020 of 1.3 million TB cases.^7^ This reduction was concentrated in 16 countries, including Brazil, which together accounted for 93% of the total global drop.^7^

The COVID-19 pandemic’s impact in the TB case notification in Brazil has been demonstrated as data become available.^6,7^ However, to our best knowledge, a few studies have estimated the unnotified cases over the two years of the pandemic and none of that assessed the State-level and group population impact on TB notification in Brazil. Moreover, prediction of expected cases should consider regional/local trends of both tuberculosis incidence and its main determinants.

In this study we estimated the unnotified cases of TB over the first two years (2020-2021) of COVID-19 pandemic in Brazil. We also calculated the difference of expected TB cases and TB case notification by sex, age, and State to identify in which population group the pandemic had the most significant impact on TB case underreporting.

## Methods

### Study design and population

This is an ecological study of new TB cases (all clinical forms) recorded in the period of 2015 to 2021 in Brazil. Brazil is a middle-income country in South America and is included in the WHO list of countries with the highest TB burden.^7^ Brazil had 212.6 million inhabitants in 2020, it occupied the 85th position of Gross Domestic Product (GDP) per capita among 195 countries and in 2017 it presented high inequality of income distribution, occupying the 8th position with the worst Gini coefficient in a list of 159 countries.^8^

### Study variables

Without patient identification, data on TB notification were obtained from the National Information System on Notifiable Diseases (Sinan).^9^ Sinan is a national system of compulsory notification of diseases of public health relevance in Brazil. The WHO estimated before the pandemic a detection rate of 87% of TB cases reported in this system.^4^

For explanatories variables, we considered its associated with the incidence of TB based on the theoretical framework of previous studies.^10^ The coverage of health services and the number of HIV-infection cases were extracted from official sources of the Brazilian Ministry of Health.^11^ The socioeconomic factors of the States were extracted from the Continuous National Household Sample Survey (Pnad).^12^ Finally, we used the Brazilian Ministry of Health estimates for the population.^13^ Table 1 describes the variables and respective data sources.

**Table 1.** Data description and source.

| Variable | Definition | Data source |
| --- | --- | --- |
| Tuberculosis case | Tuberculosis cases by sex, age group and State over 2015 to 2021 | Sinan |
| HIV-infection cases | HIV-infection cases recorded by State over 2015 to 2020 | Datasus |
| Coverage of Primary Health Care | Estimate of the population covered by Primary Health Care teams by State over 2015 to 2020 (3,000 individuals covered by one team) | e-Gestor AB |
| Coverage of Family Health Strategy | Estimate of the population covered by the Family Health Strategy teams by state over 2015 to 2020 (3,450 individuals covered by one team) | e-Gestor AB |
| Gini coefficient | Dispersion of income distribution among the members of a population by State over 2015 to 2020. A coefficient of zero indicates a perfectly equal distribution of income | Pnad |
| Average household income per capita | Household income in dollars*, in nominal terms, by total residents by State over 2015 to 2020 | Pnad |
| Unemployment rate | Percentage of people in the workforce who are unemployed by State over 2015 to 2020 | Pnad |
| Proportion of poverty | Percentage of people with a per capita household income of less than US\$5.5 per State over 2015 to 2020 | Pnad |
| Population estimate | Estimated population by State over 2015 to 2021 | Datasus |

Abbreviations: Sinan, National Information System on Notifiable Diseases; Datasus, Department of Informatics of the Health Unic System; e-Gestor AB, e-Gestor Primary Care; Pnad, Continuous National Household Sample Survey.
\*Average conversion rate of the annual quotation for the period from 2014 to 2020, R\$1/U\$3.52.

### Data analysis

We used 2015 to 2019 as COVID-19 pre-pandemic period for the TB cases prediction model. We did not included data prior to 2015 due to non-linear trends observed on the historical TB notification rate (Supplementary Material, Figure S1), possible associated to changes in socioeconomic determinants of TB in Brazil. The years 2020 and 2021 were considered as pandemic period since the first confirmed COVID-19 case in Brazil was on February 26, 2020.^14^

For each year from 2015 to 2021, TB cases data were disaggregated for population groups defined by the variables sex, age group (0-15, 15-29, 30-59, 60 and over), and State (26 States and the Federal District). The annual TB notification rate was calculated for each stratum using population estimates by sex and age group of the States. We discrebed the covariables calculating the median and interquartile range.

The TB notification rate in the pre-pandemic period (2015 to 2019) was modeled by a mixed effects generalized linear model (GLM) with Poisson regression.^15^ The equation of this model allowed for variation at the State level and in the temporal trend of TB notification by sex, age group and contextual covariates.

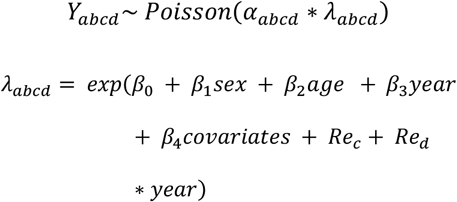

In this function, *Y*_*abcd*_ represents the count of TB cases in each stratum defined by sex (“*a*”), age group (“*b*”), State (“*c*”) and year (“*d*”), *α*_*abcd*_ represents the population size for each stratum, and *λ*_*abcd*_ represents the TB notification rate for each stratum. The equation for the *λ*_*abcd*_ includes fixed effects (*β*) for each individual variable (sex and age group), year and covariates. Furthermore, it includes the random effect (*Re*_*c*_) of States and the intracluster time trend (*Re*_*d*_ * *year*).

We first fit the null model with the random effect and added the covariates starting with the individual level (sex and age group). Contextual covariates were included in the model with one-year lag about TB case notifications because we assumed that period as latency time. The lowest Akaike criterion was used to define covariates maintenance, until all had been tested. We presented the incidence rate ratio and 95% confidence interval (95% CI).

HIV-infection detection rate, Coverage of Primary Health Care, Coverage of Family Health Strategy, unemployment rate and poverty of proportion were analyzed as continuous variables, as they presented a progressive gradient of association with the incidence of TB. The Gini coefficient was stratified into the following categories: <0.50; 0.50 to 0.55 and >0.55, as it presented a better fit with the distribution of the TB incidence rate.

Considering that the HIV-infection notification could also be impacted by the pandemic, as a sensitivity analysis we ran the model without HIV-infection detection rate as an alternative to estimate the expected TB cases.

Using these models and assuming the continuity of the trend from 2015 to 2019, we extrapolate this trend to predict TB cases in 2020 and 2021 by sex, age group and State. We used bootstrap with 1,000 simulations to estimate the standard error and the 95% uncertain intervals (95% UI) for each analytical stratum. The number of unnotified TB cases was calculated by the difference between the case notification and that predicted in 2020 and 2021 with the model. A p-value ≤ 0.05 was considered statistically significant.We also calculated the proportion of underreporting during the pandemic years regarding the expected cases according to the projection of the model obtained with the 2015-2019 data.

## Results

Between 2015 and 2021, 511,092 new TB cases were reported in Sinan, 68.7% were men, and 50.9% had 30 to 59 years age. The TB notification rate progressivily increased in the pre-pandemic period (2015-2019) from 34.3 (cases per 100,000) in 2015 to 37.1 in 2019. However, during the pandemic, the notification rate decreasead to 32.6 in 2020 and 34.0 in 2021 (Table S1).

Regarding the State level covariates, the median of the HIV-infection detection rate per 100,000 was the lowest in 2020 (14.5), and the median coverage of the Primary Health Care (82.1%), Family Health Strategy (74.1%) and unemployment rate (13.9%) was the highest in 2020 (Table S2).

In the multiple model, in addition to the demographic variables (sex and age group), the following State-level variables remained associated with the count of TB cases: HIV-infection detection rate per 100 thousand population, Gini coefficient and proportion of poverty (Table 2).

**Table 2.** Association of tuberculosis incidence cases and covariables in Brazil over the COVID-19 pre-pandemic period (2015-2019)*

| Individual level | IRR (95% CI)# | Multiple model (95% CI) |
| --- | --- | --- |
| **Age** |  |  |
| 0-14 | Reference | Reference |
| 15-29 | 8.69 (8.53-8.86) | 8.74 (8.58-8.91) |
| 30-59 | 8.77 (8.61-8.94) | 8.97 (8.8-9.14) |
| 60 and more | 8.05 (7.89-8.22) | 8.54 (8.37-8.71) |
| **Sex** |  |  |
| Women | Reference | Reference |
| Men | 2.29 (2.27-2.31) | 2.34 (2.32-2.35) |
| **State level** |  |  |
| HIV-infection detection rate per 100 thousand population\$ | 1.56 (1.32-1.85) | 1.64 (1.39-1.94) |
| Coverage of Primary Health Care (%)† | 0.95 (0.89-1.02) |  |
| Coverage of Family Health Strategy(%)† | 0.95 (0.90-1.01) |  |
| **Gini coefficient** |  |  |
| < 0.50 | Reference | Reference |
| 0.50 - 0.54 | 0.98 (0.97-1.00) | 0.98 (0.96-0.99) |
| ≥0.55 | 0.96 (0.90-1.01) | 0.95 (0.90-1.00) |
| Average household income per capita (US\$)‡ | 1.01 (0.98-1.04) | |
| Unemployment rate (%)† | 1.12 (1.05-1.20) |  |
| Proportion of poverty (%)†,§ | 1.07 (1.01-1.14) | 1.11 (1.05-1.18) |
| Year | 1.03 (1.02-1.04) | 1.03 (1.02-1.04) |

\*Mixed effects Generalized Linear Model; #For State level variable IRR adjusted by sex and age group ;\$Every 50 HIV-infection cases/100 thousand population ;†Every 20% ;‡Every 100 dollars; §Proportion of population with income less than US\$5.5/month. Abbreviations: IRR, incidence rate ratio; CI, confidence interval

Over the pre-pandemic period (2015– 2019), the number of cases and case rates estimated by the fitted model increased. The national TB case rate went from 34.1 (95% UI: 29.9, 38.3) in 2015 to 37.4 (95% UI: 32.5, 42.2) cases per 100,000 in 2019, with an average annual increase of 2.33% (95% UI: 2.10%, 2.51%) (Figure 1). TB case and case rate trends also increased in men and women (Figure S2).

**Figure 1.**
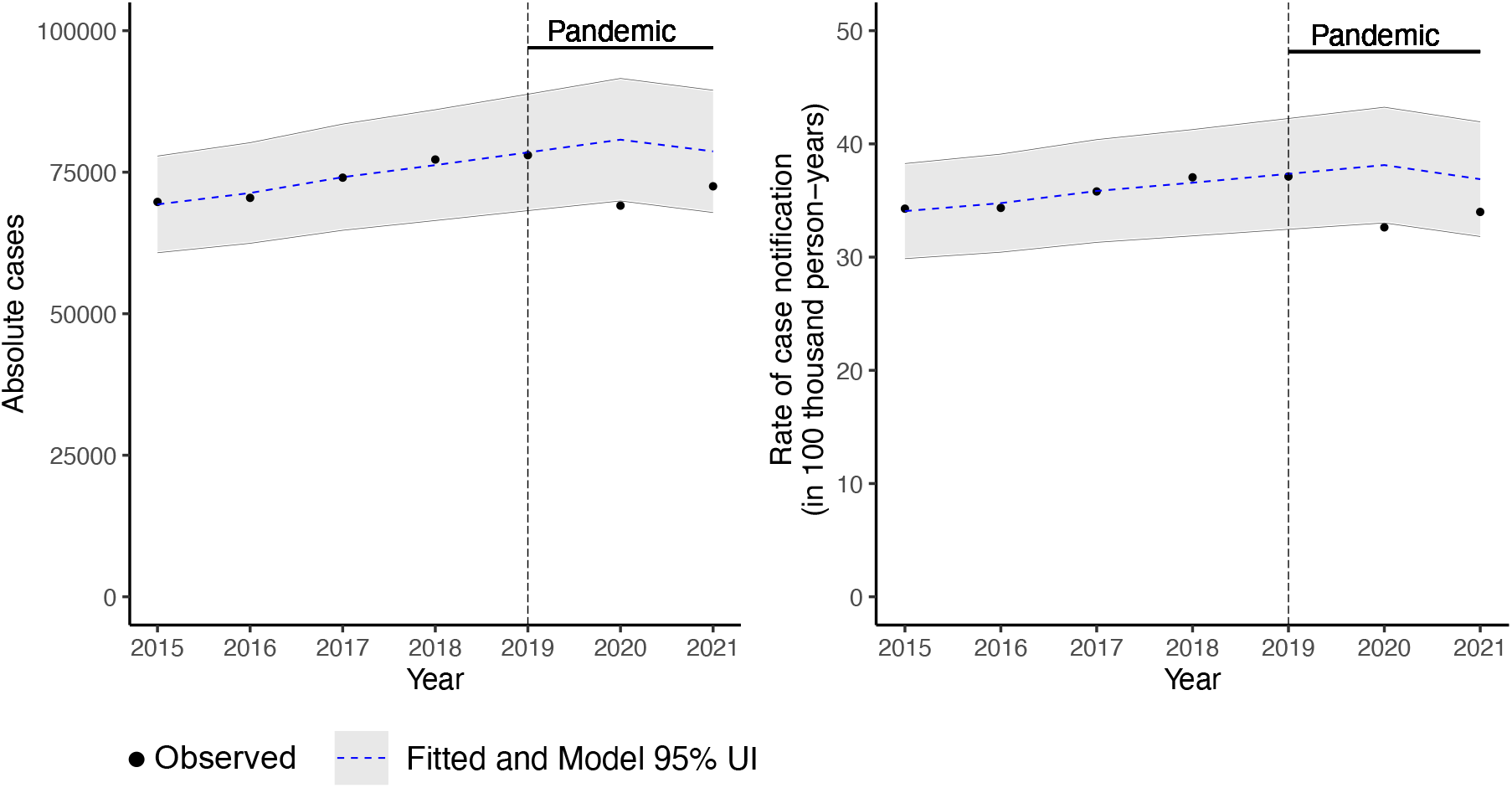
Trends in tuberculosis case notification during pre-pandemic and pandemic periods, in the overall population in Brazil over 2015 to 2021. Abbreviations: UI, Uncertain interval

Compared to 2019, the TB cases rate predicted by the model for 2020 (38.1 cases per 100,000; 95% UI: 33.0, 43.2) increased 2.08% (95% UI: 1.73%, 2.35%), and in 2021 (36.9 cases per 100,000; 95% UI: 31.8, 41.9) it decreased 1.26% (95% UI: -1.97%, -0.71%) compared to 2019 (Figure 1). Similar trend was observed in both sex (Figures S2) and most of the States (Figures S3, S4).

As expected, the national estimate of unnotified TB cases had an average of zero during the period 2015 to 2019. However, it increased significantly during 2020-2021, reaching11647 (95% UI: 829, 22466) in 2020, and 6170 (95% UI: -4629, 16968) in 2021, totaling 17817 (95% UI: -50668, 86303) unnotified cases over the two pandemic years (Table 3). Based on the expected new TB cases, we calculated 14.4% of underreporting in 2020, and 7.8% in 2021, an overall of 11.2% in both years (Table 3).

**Table 3.** Case notification, estimate of expected and unnotified tuberculosis cases* and relative underreporting in Brazil over 2015-2021.

| Years | Case notification | Estimated expected cases<br>(95% UI) | Estimate of unnotified<br>cases<br>(95% UI) | Relative<br>underreporting<br>(%)# |
| --- | --- | --- | --- | --- |
| 2015 | 69749 | 69311 (60772, 77849) | -438 (-8977, 8100) | -0.6 |
| 2016 | 70465 | 71322 (62438, 80207) | 857 (-8027, 9742) | 1.2 |
| 2017 | 74034 | 74120 (64741, 83499) | 86 (-9293, 9465) | 0.1 |
| 2018 | 77238 | 76246 (66460, 86032) | -992 (-10778, 8794) | -1.3 |
| 2019 | 78007 | 78494 (68213, 88774) | 487 (-9794, 10767) | 0.6 |
| 2020 | 69092 | 80740 (69921, 91558) | 11647 (829, 22466) | 14.4 |
| 2021 | 72507 | 78677 (67878, 89475) | 6170 (-4629, 16968) | 7.8 |
| **Total 2020-2021** | **141599** | **159416 (137800, 181033)** | **17817 (-3799, 39434)** | **11.2** |

\*Mixed effects Generalized Linear Model over the COVID-19 pre-pandemic period (2015-2019); #Percent calculated by each year as [(estimated expected cases – case notification)/ (estimated expected cases) \* 100]. Abbreviations: UI, Uncertain interval.

The absolute estimate of unnotified TB cases was substantially greater in men (2020: 7726 [95% UI: 293, 15158]; 2021: 4083 [95% UI: -3335, 11500]) compared to women (2020: 3922 [95% UI: 536, 7308]; 2021: 2087 [95% UI: -1294, 5468]) (Table 4, Figure S5). Across sex and age strata, men aged 30 to 59 years had the highest number of unnotified cases (2020: 3906 [95% UI: 89, 7723]; 2021: 1933 [95% UI: -1890, 5756]). The greater impact in the relative underreporting was in men with 0 to 14 years of age. In 2020, 40.5% of expected TB cases in this group were not notified, and in 2021, 29.4% (Table 4, Figure S5).

**Table 4.** Estimate of unnotified tuberculosis cases* and relative underreporting by sex and age group in Brazil over the COVID-19 pandemic period 2020-2021.

| Sex/Age<br>group<br>(years) | 2020 |  | 2021 |  | Total 2020-2021 |  |
| --- | --- | --- | --- | --- | --- | --- |
|  | Estimate of<br>unnotified cases<br>(95% UI) | Relative<br>underreporting<br>(%)# | Estimate of<br>unnotified cases<br>(95% UI) | Relative<br>underreporting<br>(%)# | Estimate of<br>unnotified cases<br>(95% UI) | Relative<br>underreporting<br>(%)# |
| **Women** | 3922 (536, 7308) | 15.5 | 2087 (-1294, 5468) | 8.4 | 6009 (-758, 12776) | 12.0 |
| **Age** |  |  |  |  |  |  |
| 0-14 | -193 (-288, -98) | -27.3 | -369 (-462, -276) | -54.4 | -562 (-750, -374) | -40.5 |
| 15-29 | 669 (-275, 1613) | 9.5 | 68 (-853, 989) | 1.0 | 737 (-1128, 2602) | 5.4 |
| 30-59 | 2377 (644, 4109) | 18.3 | 1690 (-43, 3423) | 13.4 | 4067 (601, 7533) | 15.9 |
| 60 and more | 1069 (455, 1684) | 22.9 | 698 (63, 1332) | 14.9 | 1767 (518, 3015) | 18.9 |
| **Men** | 7726 (293, 15158) | 13.9 | 4083 (-3335, 11500) | 7.6 | 11808 (-3041, 26658) | 10.8 |
| **Age** |  |  |  |  |  |  |
| 0-14 | 700 (468, 933) | 40.5 | 488 (260, 715) | 29.4 | 1188 (728, 1648) | 35.1 |
| 15-29 | 1752 (-497, 4000) | 10.5 | 915 (-1283, 3112) | 5.7 | 2667 (-1779, 7113) | 8.2 |
| 30-59 | 3906 (89, 7723) | 13.7 | 1933 (-1890, 5756) | 6.9 | 5839 (-1801, 13478) | 10.4 |
| 60 and more | 1367 (233, 2502) | 16.1 | 748 (-421, 1917) | 8.8 | 2115 (-188, 4419) | 12.4 |
| **Total** | 11647 (829, 22466) | 14.4 | 6170 (-4629, 16968) | 7.8 | 17817 (-3799, 39434) | 11.2 |

\*Mixed effects Generalized Linear Model over the COVID-19 pre-pandemic period (2015-2019); # Percent calculated by each strata as: [(estimated expected cases – case notification)/ (estimated expected cases) \* 100]. Abbreviations: UI, Uncertain interval.

The number of unnotified cases varied greatly by States, and it was significant for 13 States of the 27 (48.1%) in 2020, and for 4 (14.8%) in 2021 (Table 5, Figure S6). The highest number of unnotified cases was high in São Paulo (2514; 95% UI: 328, 4701) and Rio de Janeiro (1778; 95% UI: 875, 2680) in 2020; and in São Paulo (2446; 95% UI: 160, 4732) and Rio Grande Sul (961; 95% UI: 350, 1572) in 2021 (Table 5).

**Table 5.** Estimate of unnotified tuberculosis cases* and relative underreporting by States in Brazil over the COVID-19 pandemic period 2020-2021.

| State | 2020 |  | 2021 |  | Total 2020-2021 |  |
| --- | --- | --- | --- | --- | --- | --- |
|  | Estimate of unnotified cases (95% UI) | Relative underreporting (%)† | Estimate of unnotified cases (95% UI) | Relative underreporting (%)† | Estimate of undetected cases (95% UI) | Relative unnotified (%)† |
| **North** | 1629 (524, 2734)<sup>#</sup> | 15.8 | 206 (-793, 1205) | 2.1 | 1835 (-269, 3939) | 9.2 |
| Acre | -13 (-56, 31) | -2.5 | 3 (-40, 46) | 0.6 | -10 (-96, 77) | -0.9 |
| Amazonas | 836 (548, 1125)<sup>#</sup> | 23.0 | 371 (112, 630)<sup>#</sup> | 10.7 | 1207 (659, 1754)<sup>#</sup> | 17.0 |
| Amapá | -5 (-52, 41) | -2.1 | -73 (-119, -27)<sup>#</sup> | -28.0 | -79 (-171, 14) | -15.0 |
| Pará | 659 (136, 1182)<sup>#</sup> | 13.8 | -51 (-502, 401) | -1.2 | 608 (-366, 1582) | 6.7 |
| Rondônia | 125 (40, 210)<sup>#</sup> | 21.0 | 40 (-48, 128) | 6.8 | 165 (-8, 337) | 14.0 |
| Roraima | 14 (-31, 59) | 4.4 | -42 (-78, -7)<sup>#</sup> | -14.9 | -28 (-109, 52) | -4.7 |
| Tocantins | 13 (-61, 88) | 6.7 | -41 (-117, 35) | -21.0 | -28 (-178, 122) | -7.0 |
| **Northeast** | 3147 (52, 6241)<sup>#</sup> | 15.1 | 990 (-2009, 3990) | 5.0 | 4137 (-1957, 10231) | 10.2 |
| Alagoas | 221 (37, 405)<sup>#</sup> | 21.3 | 135 (-45, 314) | 13.5 | 355 (-8, 719) | 17.5 |
| Bahia | 776 (0, 1551)<sup>#</sup> | 17.3 | 111 (-647, 869) | 2.7 | 887 (-647, 2421) | 10.2 |
| Ceará | 713 (225, 1202)<sup>#</sup> | 19.0 | 427 (-61, 914) | 11.6 | 1140 (164, 2116)<sup>#</sup> | 15.3 |
| Maranhão | 204 (-214, 623) | 8.9 | -244 (-628, 139) | -11.7 | -40 (-842, 762) | -0.9 |
| Paraíba | 240 (25, 455)<sup>#</sup> | 18.9 | 120 (-93, 332) | 9.7 | 360 (-68, 788) | 14.3 |
| Pernambuco | 789 (261, 1318)<sup>#</sup> | 15.7 | 147 (-368, 662) | 3.1 | 936 (-107, 1980) | 9.5 |
| Piauí | 94 (-77, 265) | 12.7 | -2 (-165, 161) | -0.3 | 92 (-243, 426) | 6.4 |
| Rio Grande do Norte | -26 (-213, 161) | -2.0 | 177 (-3, 358) | 14.0 | 151 (-216, 518) | 5.8 |
| Sergipe | 135 (10, 260)<sup>#</sup> | 15.7 | 121 (0, 241)<sup>#</sup> | 14.2 | 256 (10, 501)<sup>#</sup> | 15.0 |
| **Southeast** | 4663 (392, 8934)<sup>#</sup> | 12.9 | 3150 (-1276, 7576) | 8.7 | 7813 (-883, 16510) | 10.8 |
| Espírito Santo | -54 (-249, 140) | -4.3 | -276 (-477, -75)<sup>#</sup> | -22.5 | -330 (-725, 65) | -13.2 |
| Minas Gerais | 425 (-563, 1413) | 11.5 | 324 (-710, 1359) | 8.9 | 750 (-1273, 2772) | 10.2 |
| Rio de Janeiro | 1778 (875, 2680)<sup>#</sup> | 14.1 | 656 (-249, 1560) | 5.2 | 2433 (627, 4240)<sup>#</sup> | 9.7 |
| São Paulo | 2514 (328, 4701)<sup>#</sup> | 13.6 | 2446 (160, 4732)<sup>#</sup> | 13.1 | 4961 (488, 9433)<sup>#</sup> | 13.4 |
| **South** | 1609 (56, 3161)<sup>#</sup> | 16.9 | 1381 (-178, 2939) | 14.7 | 2989 (-122, 6101) | 15.8 |
| Paraná | 183 (-361, 727) | 7.8 | 304 (-257, 865) | 13.1 | 487 (-617, 1592) | 10.5 |
| Rio Grande do Sul | 1056 (433, 1680)<sup>#</sup> | 19.8 | 961 (350, 1572)<sup>#</sup> | 18.3 | 2017 (783, 3252)<sup>#</sup> | 19.0 |
| Santa Catarina | 369 (-16, 754) | 20.0 | 116 (-271, 502) | 6.4 | 485 (-287, 1257) | 13.3 |
| **Midwest** | 601 (-195, 1396) | 15.2 | 442 (-373, 1258) | 11.4 | 1043 (-568, 2654) | 13.4 |
| Distrito Federal | 67 (-80, 214) | 18.3 | 58 (-96, 212) | 15.6 | 125 (-176, 426) | 17.0 |
| Goiás | 132 (-199, 463) | 12.6 | 80 (-267, 426) | 7.7 | 212 (-466, 889) | 10.2 |
| Mato Grosso do Sul | 178 (39, 318)<sup>#</sup> | 14.5 | 80 (-57, 217) | 6.6 | 258 (-18, 535) | 10.6 |
| Mato Grosso | 223 (46, 401)<sup>#</sup> | 17.2 | 225 (46, 403)<sup>#</sup> | 17.8 | 448 (92, 804)<sup>#</sup> | 17.5 |
| **Total** | 11647 (829, 22466)<sup>#</sup> | 14.4 | 6170 (-4629, 16968) | 7.8 | 17817 (-3799, 39434) | 11.2 |

\*Mixed effects Generalized Linear Model over the COVID-19 pre-pandemic period (2015-2019); †Percent calculated by each locality as: [(estimated expected cases – case notification)/ (estimated expected cases) \* 100] #P<0.05. Abbreviations: UI, Uncertain interval.

In 2020, 23.0% of expected TB cases in Amazonas were underreported, and in 2021, 10.7%. In Alagoas, 21.3% and 13.5% of TB cases were not reported, respectively in 2020 and 2021. Rondônia also presented a high relative underreporting of cases, with 21.0% in 2020 and 6.8% in 2021 (Table 5, Figure S6).

### Sensitivity analysis

In the sensitivity analysis without HIV-infection detection rate, we estimated 11290 (95% UI: -255, 11545) underreported TB cases in 2020, 8866 (95% UI: -3100,11966) in 2021, and a total of 20156 (95% UI: 53163, 73319) in both years (Table S3). This was 2338 more unnotified TB cases than the main model (Table 3). The overall estimated underreporting TB cases for 2020 was 14.0% and similar to the main model (14.4%). However, for 2021, the model without HIV-infection detections rate estimated 10.9% of underreporting cases (Table S3), higher than that obtained in the main analysis (7.8%) (Table 3).

## Discussion

In this study, we estimated the underreporting of TB cases in Brazil during the first two years of the COVID-19 pandemic. Except for women aged 0 to 14 years, all population groups presented an absolute reduction of TB case notifications, with the greatest impact on men aged 30-59 years. The group population with greater impact in relative underreporting of TB cases was men with 0 to 14 years age. Undernotification of cases varied widely between States, with the greatest relative underreporting estimated for Amazonas in 2020.

The impact on national-level TB notifications in 2020 (14.4%) was similar to that reported in Mozambique (15.1%)^3^, but lower than in India, that had a relative loss of cases of 63.3% during the period from March 2020 through April 2021.^16^ Other studies estimated the effect of the pandemic on TB case notifications with different approaches and also have found an immediate impact. Reported data from facilities in 2020 revealed a decline in TB case notifications of 34%^17^ in Nigeria and of 43% in Uganda.^18^ In addition, WHO estimates suggests that the countries that contributed most to the global shortfall in TB notifications between 2019 and 2020 were India (41%), Indonesia (14%), the Philippines (12%) and China (8%).^7^

There are numerous explanations for the TB case underreport. Reallocation of resources in response of the pandemic severely disrupted the provision of TB health services because the entire diagnostic and health care network focused on pandemic response. This affected the access of appropriate equipment and capacity, commodities and stock. In addition, fearing of SARS-CoV-2 infection and stigma, restrictions to movement and reduced service uptime limited patients’ access to health services.^19^ And finally, active case finding were discontinued in community and household.^1,2^

We also observed that although the national-level underreport in TB cases remained in 2021, it was smaller than 2020 and not significant. Several factors may have contributed to this. One could be the result of efforts by federal management, local authorities, and health services to retake TB control activities, that may have taken effect later, in 2021. Right at the beginning of the pandemic in Brazil, federal guidelines to assist local TB programmes and health services were published defining TB as a differential diagnosis of COVID-19 and as a risk factor for complications of COVID-19 outcomes.^20^ Considering the worldwide reducing of TB case reporting, WHO also released recommendations for national TB programmes and health personnel to urgently maintain continuity of essential services for people affected with TB during the COVID-19 pandemic.^21^ Another factor that may have contributed to this was the availability of the vaccine against SARS-CoV-2 in 2021. The immunization of the population resulted, not only in Brazil, but in the world, in a progressive flexibilization of the social distance measures imposed during the pandemic, favoring the seeking of the population for services for health problems other than COVID-19.^22^

Finally, although a decrease in HIV-infection cases has been observed in almost the entire country, especially in recent years, HIV-infection case detection rate, which was a covariate in the model, was also impacted by the pandemic.^23^ When compared to 2019, the HIV-infection detection rate decreased 16.1% in 2020. Because predictive variables were analysed with one-year lag in relation to TB case notifications, potential underreporting of HIV-infection cases in 2020 could underestimate the predictions of TB cases in 2021, which means that the relative underreporting of TB cases could be greater for this year. To assess the effect of underreporting of HIV-infection cases, we run a sensitivity analysis without this covariable that estimated an adittion of 2696 unnotified TB cases in 2021. This indicates that the underreporting in 2021 could be larger than the one calculated in the main model, but still smaller than the one in 2020.

The concentration of underreporting of TB cases in men is consistent with the high risk of TB in this group.^24^ In Brazil, they are also the most affected by unfavorable treatment outcomes^25^, which might be an explanation for the greater impact of the pandemic. Furthermore, the majority of the prison population in Brazil are young men, and although the number of TB cases in this population had been increasing in the country, a Brazilian TB Bulletin launched in the TB World Day in 2022 reported a decrease of TB case notification in prison population during the pandemic years^26^, suggesting that TB case finding in this population may also have suffered the impacts of the pandemic.

Similar to ours findings, in the first year of the pandemic a study in Mozambique found that men were the most affected in terms of TB notification, resulting in a 15% (95% CI 4.0 to 25.0) relative loss, and suggested that this was due to the privileges that women and children had in the country to access health services.^3^ On the other hand, a study in Zambia found that the proportional distribution of TB notifications according to sex did not significantly differ over the pandemic period.^27^ Considering that just a few studies analyzed the pandemic COVID-19 impact on TB case notification by population group, the understanding of this association requires further investigation.

Our estimates showed a higher relative underreporting of cases in men aged 0 to 14 years. The diagnosis of TB in children is challenging due to insufficient specimen material and the scarcity of bacilli in specimens.^28^ In these situations, invasive procedures such as nasopharyngeal aspirate and gastric lavage are indicated. Moreover, symptoms in children are generally nonspecific and are confused with childhood infections, which makes assessment difficult.^28^ These difficulties may have been exacerbated by the disorganization in services that the pandemic caused. Moreover, since children are usually contacted from sick adults, the underdetection of TB cases in adults, and discontinuation of the contact tracing could also be an explanation.^1^

During the pandemic, the State of Amazonas was classified as one of the most fragile States with regard to the capacity to serve the population, especially in critical situations, with high demand shock.^29^ In addition, it consistently presented the highest incidence rate of COVID-19.^30^ These factors may explain the greater underreporting of cases that we found in this State.

We consider that our study has some limitation. The national TB case notifications is affected by case detection even before the pandemic^31^, which make it difficult to isolate the cause of the observed trend. This could also be a limitation for the estimates by States and group population. In addition, because of non-availability of data, it was not possible to analyze trends of TB case notifications across others population subgroups, as for example race/ethnicity, which could help us to understand better the impact of the pandemic in TB cases by social disparities. Finally, the non-pharmacological interventions adopded during the pandemic may lead to an eventual reduction in TB transmission, which could partly explain the reduction in notifications. We did not have information to quantify this potential phenomenon. However, if this reduction in TB transmission were also observed in other infectious diseases, the reduction in HIV cases could help to model this trend. Based on this, we considered the model with HIV as the main model to obtain a more conservative measure of the underreporting estimate.

While what we observed in 2021 suggests a possible overcome of the negative effects of COVID-19 disruptions in TB health services in Brazil, the pandemic left 17817 people without TB diagnosis in two years (may be more according to the sensitivity analysis). It is uncertain of what impact those undetected cases will represent in the future epidemiology of TB. Modeling estimates from early in the pandemic revealed an immediate increase in TB mortality^32^ which was not seem in Brazil.^26^ Misclassification of cause of deaths could be an explanation. Confinement can cause long-lasting increases in TB burden due intra-household transmission of undiagnosed cases,^32^ which would be expected in the next years.

The catastrophic effect of COVID-19 pandemic in Brazil, resulted in a setback in progress made over decades in TB control and a potential delay in achieving the 2030 targets of the WHO End TB Strategy. Several lessons learnt from COVID-19 responses are an opportunity to improve the detection of respiratory diseases.1 In this way, we identify the populational groups particularly affected by the underreporting of TB. New ways of working, integrated services to use existing resources maximally, and patient-centered tuberculosis service, could have a synergistic, enhancing, and multiplier effect in restoring TB control activities.

## Data Availability

All data used in this study are openly accessible and available through the sources listed in the manuscript.

## Contributors statement

DMP, PBO and FADQ conceptualized the study. DMP, PBO, FDCJ and FADQ contributed to study design. DMP curated data. DMP and FADQ contributed methodology, formal analysis and validation. DMP drafted the first version of the Article. DMP, PBO, FDCJ and FADQ reviewed and edited the manuscript. All authors read and met the ICMJE criteria for authorship and agree with the results and conclusions. All authors had full access to all the data in the study and had final responsibility for the decision to submit for publication.

## Declaration of interests

The authors declare no conflicts of interest.

## Funding

FADQ is beneficiary of a fellowship for research productivity from the National Council for Scientific and Technological Development - CNPq: 312656/2019-0. This study was financed in part by the Coordenação de Aperfeiçoamento de Pessoal de Nível Superior - Brazil CAPES tese award (Edital Nº 3/2021).

## Notes

### Competing Interest Statement

The authors have declared no competing interest.

