## Supplemental material for "Impact of the COVID-19 pandemic on tuberculosis notification in Brazil"

**TABLES**

**Table S1. Number and tuberculosis notification rate per 100 thousand person-year by sex and age group in Brazil over the COVID-19 pre-pandemic (2015-2019) and pandemic periods (2020-2021)**

| **Individual level** | **Number of tuberculosis cases (notification rate per 100 thousand person-year)** | | | | | | |
| --- | --- | --- | --- | --- | --- | --- | --- |
|  | **2015** | **2016** | **2017** | **2018** | **2019** | **2020** | **2021** |
| **Age (year)** |  |  |  |  |  |  |  |
| 0-14 | 2106 (4.6) | 2147 (4.8) | 2238 (5.0) | 2504 (5.6) | 2605 (5.9) | 1931 (4.4) | 2218 (5.0) |
| 15-29 | 20889 (40.4) | 21235 (41.2) | 23079 (45.1) | 24239 (47.7) | 24486 (48.6) | 21310 (42.6) | 21683 (43.7) |
| 30-59 | 36500 (44.9) | 36448 (44.1) | 37414 (44.6) | 38862 (45.6) | 38870 (45.0) | 35149 (40.2) | 36845 (41.7) |
| 60 and more | 10254 (41.0) | 10635 (40.9) | 11303 (41.9) | 11633 (41.5) | 12046 (41.4) | 10702 (35.4) | 11761 (37.5) |
| **Sex** |  |  |  |  |  |  |  |
| Women | 22302 (21.5) | 22446 (21.4) | 22864 (21.6) | 24030 (22.6) | 24411 (22.7) | 21434 (19.8) | 22631 (20.8) |
| Men | 47447 (47.6) | 48019 (47.8) | 51170 (50.6) | 53208 (52.2) | 53596 (52.2) | 47658 (46.0) | 49876 (47.8) |
| **Total** | **69749 (34.3)** | **70465 (34.3)** | **74034 (35.8)** | **77238 (37.0)** | **78007 (37.1)** | **69092 (32.6)** | **72507 (34.0)** |

**Table S2. Median and interquartile range of State level covariates* in Brazil over 2014-2020**

| **State level** | **Median (interquartile range 25%-75%)** | | | | | | |
| --- | --- | --- | --- | --- | --- | --- | --- |
|  | **2014** | **2015** | **2016** | **2017** | **2018** | **2019** | **2020** |
| Aids detection rate per 100 thousand population | 19.8  (14.7-24.8) | 18.6  (14.9-21.8) | 17.2  (14.9-23.9) | 18.7  (15.0-24.0) | 18.1  (15.0-23.8) | 17.3  (15.1-23.9) | 14.5  (12-18.5) |
| Coverage of Primary Health Care (%) | 76.9  (70.9-87.3) | 77.5  (72.1-87.5) | 78.7  (71.3-87.3) | 75.5  (71.3-86.3) | 80.6  (73.0-87.0) | 76.9  (73.2-86.1) | 82.1  (75.4-87.2) |
| Coverage of Family Health Strategy (%) | 71.1  (59.9-76.3) | 71.2  (62.0-79.3) | 73.3  (61.1-79.7) | 70.8  (56.5-79.7) | 70.4  (62.8-79.8) | 70.8  (60.4-78.9) | 74.1  (62.5-79.9) |
| Gini coefficient | 0.51  (0.48-0.53) | 0.51  (0.48-0.53) | 0.52  (0.48-0.54) | 0.53  (0.49-0.55) | 0.54  (0.49-0.55) | 0.53  (0.49-0.56) | 0.50  (0.47-0.53) |
| Average household income per capita (US$) | 217.3  (195.6-299.0) | 234.4  (213.8-313.4) | 256.0  (217.9-348.2) | 266.2  (238.5-364.8) | 296.9  (244.3-401.3) | 300  (256.1-424.9) | 301.1  (254.3-410.4) |
| Unemployment rate (%) | 7.1  (5.7-8.5) | 8.6  (7.4-9.5) | 11.5  (9.4-12.8) | 12.9  (10.2-14.4) | 12.3  (9.9-14.2) | 12.5  (10.3-14.6) | 13.9  (11-15.8) |
| Proportion of poverty (%)# | 33.2  (16.8-41.3) | 35  (17.2-41) | 35.2  (19.2-44.8) | 35.5  (18.4-45.9) | 33.5  (18.2-44.4) | 39.6  (17.1-44.4) | 34.2  (17.1-40.3) |

*Variables included in the multiple model with one year lag.

#Proportion of population with income less than US$5.5/month

**Table S3. Sensitivity analysis - Case notification, estimate of expected and unnotified tuberculosis cases* and relative underreporting in Brazil over 2015-2021**

| **Years** | **Case notification** | **Estimated expected cases  (95% UI)** | **Estimate of unnotified cases (95% UI)** | **Relative underreporting (%)#** |
| --- | --- | --- | --- | --- |
| 2015 | 69749 | 69200 (60172, 78228) | -549 (-9577, 9028) | -0.8 |
| 2016 | 70465 | 71424 (62007, 80841) | 959 (-8458, 9417) | 1.3 |
| 2017 | 74034 | 74148 (64186, 84111) | 114 (-9848, 9962) | 0.2 |
| 2018 | 77238 | 76330 (65892, 86769) | -908 (-11346, 10439) | -1.2 |
| 2019 | 78007 | 78391 (67429, 89353) | 384 (-10578, 10962) | 0.5 |
| 2020 | 69092 | 80382 (68837, 91927) | 11290 (-255, 11545) | 14.0 |
| 2021 | 72507 | 81373 (69407, 93340) | 8866 (-3100, 11966) | 10.9 |
| **Total 2020-2021** | **141599** | **161755 (138245, 185266)** | **20156 (-3354, 23511)** | **12.5** |

*Mixed effects Generalized Linear Model over the COVID-19 pre-pandemic period (2015-2019) without the covariable Aids detection rate.

#Percent calculated by each year as [(estimated expected cases – case notification)/ (estimated expected cases) * 100]

Abbreviations: UI, Uncertain interval

**FIGURES**

|  | Brasil |
| --- | --- |
| 2008 | 38.2273 |
| 2009 | 37.5716 |
| 2010 | 36.3871 |
| 2011 | 37.0202 |
| 2012 | 35.8662 |
| 2013 | 35.5953 |
| 2014 | 34.7213 |
| 2015 | 34.3083 |
| 2016 | 34.3733 |
| 2017 | 35.8294 |
| 2018 | 37.0834 |
| 2019 | 37.1625 |

**Figure S1. Tuberculosis notification rate per 100 thousand person-year in Brazil, 2008 to 2019.**

Source: National Information System on Notifiable Diseases (Sinan) and Department of Informatics of the Health Unic System (Datasus).

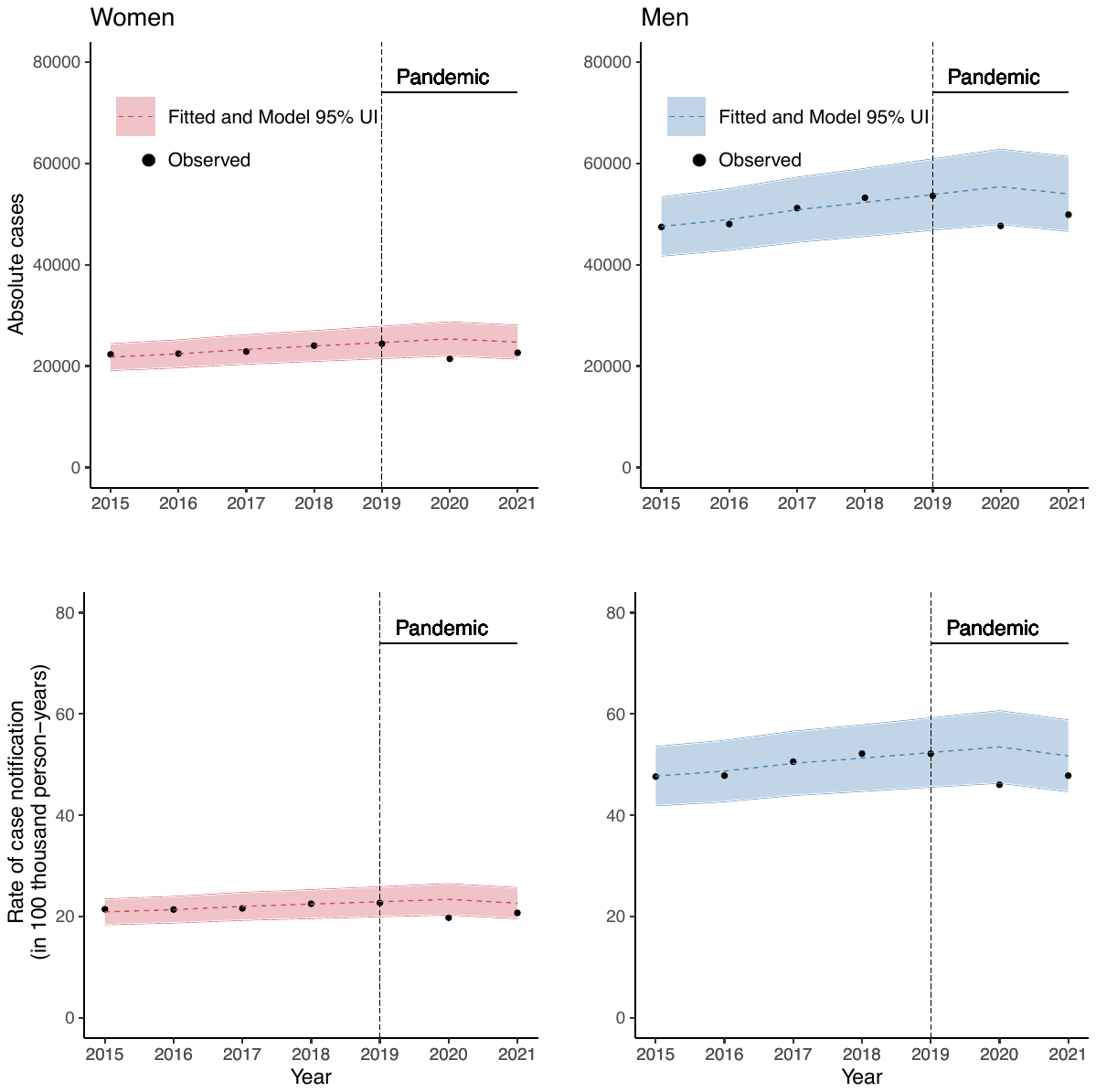

**Figure S2. Trends in tuberculosis case notification during pre-pandemic and pandemic periods by sex in Brazil over 2015 to 2021.**

Abbreviations: UI, Uncertain interval

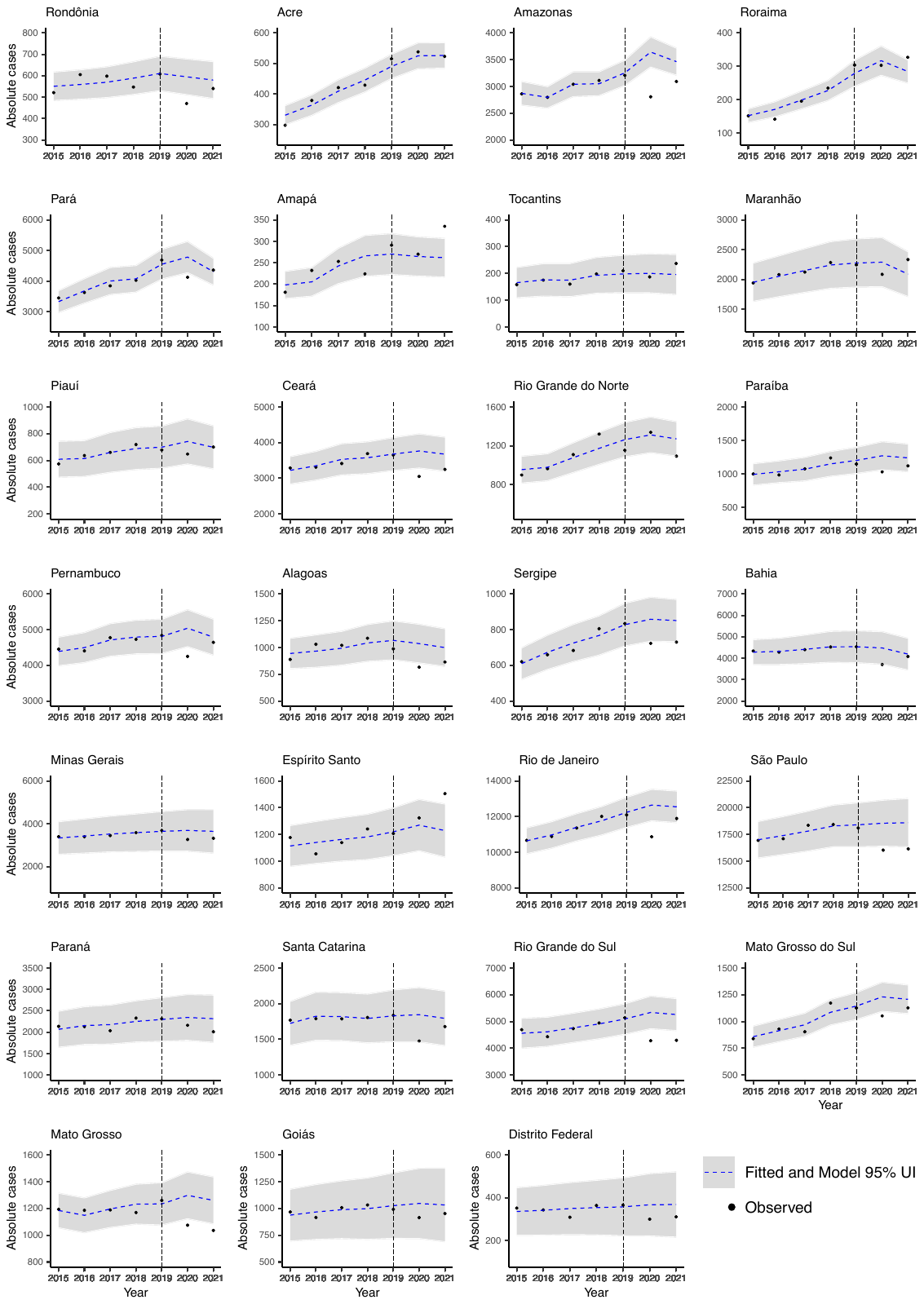

**Figure S3. Trends in the number of tuberculosis case notification during pre-pandemic and pandemic periods by State in Brazil over 2015 to 2021.**

Abbreviations: UI, Uncertain interval

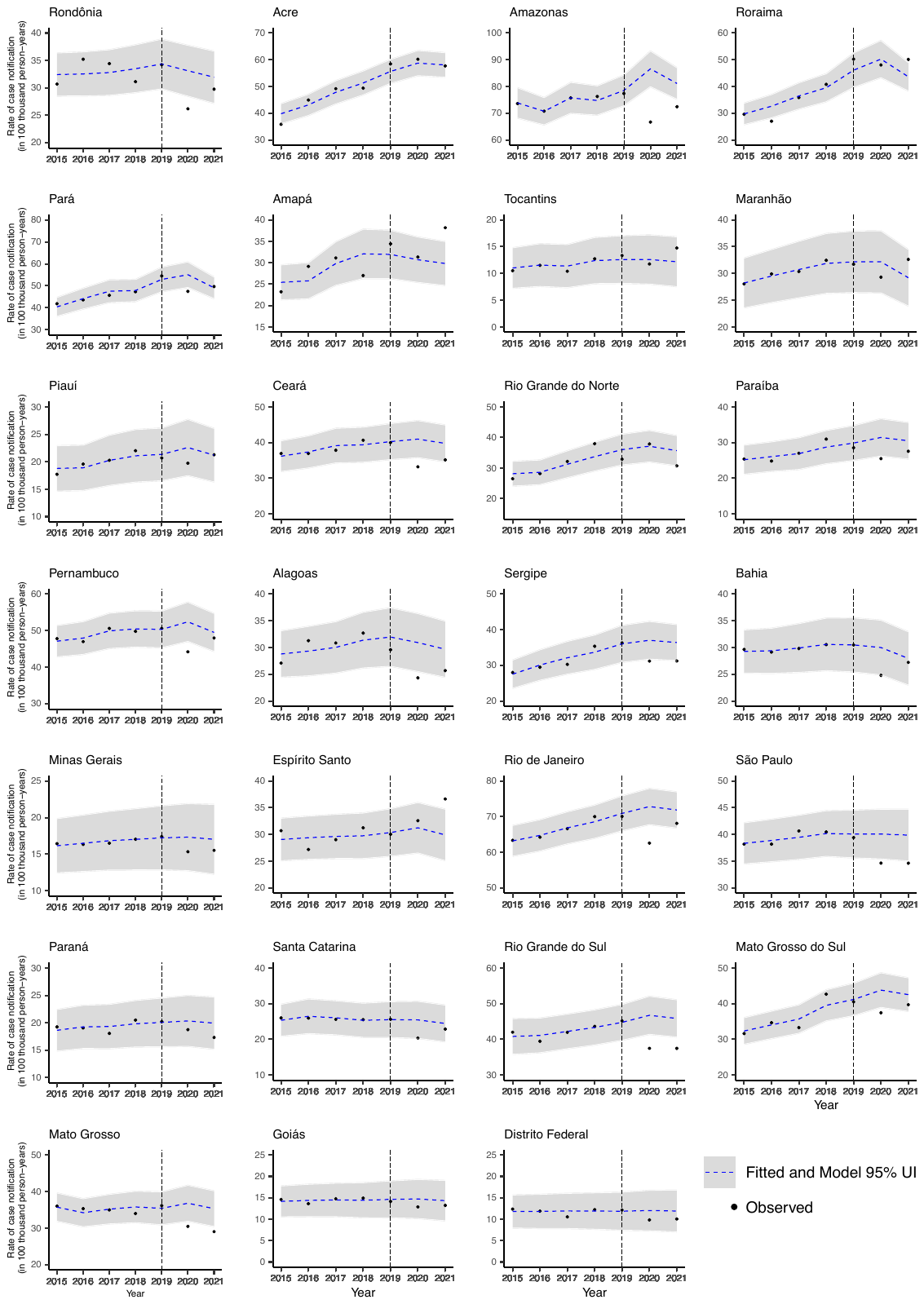

**Figure S4. Trends in tuberculosis case notification rate during pre-pandemic and pandemic periods by State in Brazil over 2015 to 2021.**

Abbreviations: UI, Uncertain interval

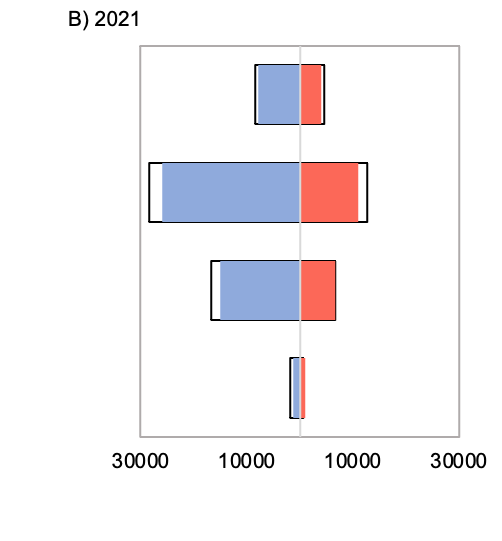

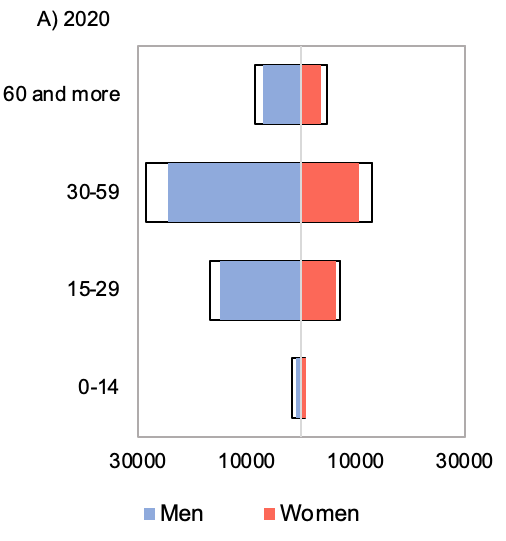

|  | Women | Men | Women | Men |
| --- | --- | --- | --- | --- |
| 0-14 | 678 | -1730.39 | 1047 | -1171 |
| 15-29 | 6697 | -16708.9 | 6629 | -15054 |
| 30-59 | 12654 | -28465.9 | 10964 | -25881 |
| 60 and more | 4689 | -8478.34 | 3991 | -7770 |

|  | Women | Men | Women | Men |
| --- | --- | --- | --- | --- |
| 0-14 | 707.855 | -1730.39 | 901 | -1030 |
| 15-29 | 7022 | -16708.9 | 6353 | -14957 |
| 30-59 | 12965.9 | -28465.9 | 10589 | -24560 |
| 60 and more | 4660.31 | -8478.34 | 3591 | -7111 |

**Figure S5. Expected tuberculosis cases (black outline) and case notifications by sex and age group in Brazil over the COVID-19 pandemic period 2020-2021**

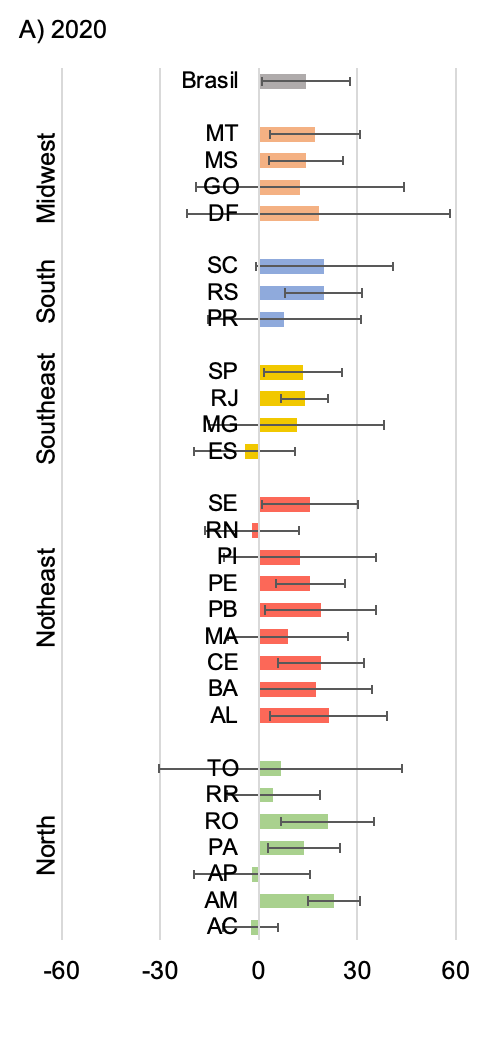

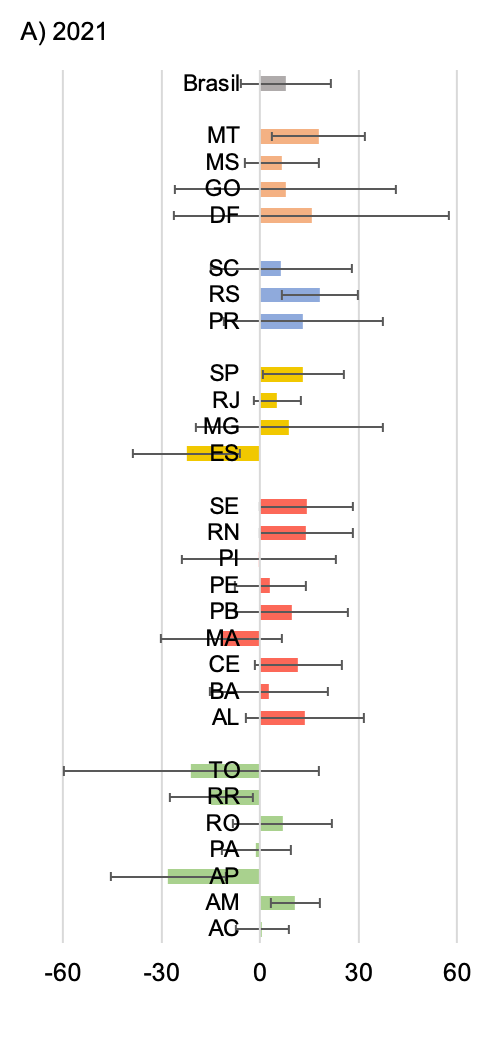

Estimated underreporting (%)

|  | 2020 |
| --- | --- |
| AC | -2.45337 |
| AM | 22.9572 |
| AP | -2.0634 |
| PA | 13.773 |
| RO | 20.9906 |
| RR | 4.3802 |
| TO | 6.6789 |
| AL | 21.3231 |
| BA | 17.329 |
| CE | 18.9514 |
| MA | 8.93243 |
| PB | 18.8996 |
| PE | 15.6636 |
| PI | 12.6519 |
| RN | -1.99668 |
| SE | 15.7379 |
| ES | -4.26923 |
| MG | 11.5285 |
| RJ | 14.0557 |
| SP | 13.5548 |
| PR | 7.81602 |
| RS | 19.7917 |
| SC | 20.0066 |
| DF | 18.2955 |
| GO | 12.6064 |
| MS | 14.4858 |
| MT | 17.1919 |
| Brasil | 14.4262 |

|  | 2020 |
| --- | --- |
| AC | 0.610945 |
| AM | 10.7045 |
| AP | -28.0231 |
| PA | -1.17489 |
| RO | 6.84061 |
| RR | -14.8698 |
| TO | -21.0328 |
| AL | 13.4746 |
| BA | 2.65461 |
| CE | 11.6049 |
| MA | -11.7049 |
| PB | 9.65899 |
| PE | 3.07091 |
| PI | -0.294203 |
| RN | 13.95 |
| SE | 14.1846 |
| ES | -22.4514 |
| MG | 8.89806 |
| RJ | 5.22582 |
| SP | 13.15 |
| PR | 13.1405 |
| RS | 18.2748 |
| SC | 6.44804 |
| DF | 15.6321 |
| GO | 7.73193 |
| MS | 6.64584 |
| MT | 17.8187 |
| Brasil | 7.84194 |

**Figure S6. Proportion and uncertain interval of estimated underreporting* tuberculosis cases by States in** **Brazil over the COVID-19 pandemic period 2020-2021**

*Percent calculated by each locality as: [(estimated expected cases – case notification)/ (estimated expected cases) * 100]

Abbreviations: AC, Acre; AM, Amazonas; AP, Amapá; PA, Pará; RO, Rondônia; RR, Roraima; TO, Tocantins; AL, Alagoas; BA, Bahia; CE, Ceará; MA, Maranhão; PB, Paraíba; PE, Pernambuco; PI, Piauí; RN, Rio Grande do Norte; SE, Sergipe; ES, Espírito Santo; MG, Minas Gerais; RJ, Rio de Janeiro; SP, São Paulo; PR, Paraná; RS, Rio Grande do Sul; SC, Santa Catarina; DF, Distrito Federal; GO, Goiás; MS, Mato Grosso do Sul; MT, Mato Grosso
